# Role of Probiotics on Dialysis Patients in End-Stage Kidney Disease: A Systematic Review

**DOI:** 10.1101/2024.12.20.24319427

**Authors:** Zaher Ahmed, Md. Sajjadul Karim

**Affiliations:** Department of Pharmacy, University of Asia Pacific, Dhaka, Bangladesh

**Keywords:** probiotics, dialysis, end-stage kidney patients, gut dysbiosis, uremic toxins

## Abstract

The prevalence of chronic kidney disease (CKD) has been increasing all over the world due to the high-risk factors of metabolic syndrome. But, unfortunately, the cost of dialysis and the scarcity of dialysis center and dialysate are making it more complex for the people in least developed countries. Probiotics are being studied as a potential treatment option for chronic kidney disease, especially in the dialysis patients. The aim of this review is to investigate the effects of probiotics administration in dialysis patients in end-stage kidney disease. A systemic search was conducted on MEDLINE database from 2002 to 2023 using key terms related to dialysis, end-stage kidney disease and probiotics. Fifteen studies met eligibility criteria, among which thirteen were on hemodialysis patients and others on peritoneal dialysis patients. The results of the studies revealed that probiotics have some significant effect on gut dysbiosis, gastrointestinal symptoms, uremic toxins, inflammation and overall quality of life of dialytic patients. Studies showed that administration of probiotics inhibit the growth of pathogenic bacteria as well as production of protein-bound uremic toxins (i.e. indoxyl sulfate and p-cresol sulfate) which can not be fully excreted by dialysis. The level of serum TNF-α, IL-5 and IL-6 were significantly decreased in peritoneal dialysis patients. However, further investigations must be carried out with larger sample size with larger study duration and wit different probiotics or synbiotics preparations to obtained more specific explanations of the effects and mechanisms of probiotics to counteract the disease progression on dialysis patients in end-stage kidney disease.

## INTRODUCTION

Chronic kidney disease (CKD) has become one of the major cause of deaths in 21^st^ century. Due to the high risk factors of metabolic syndrome, such as diabetes mellitus, high blood pressure (hypertension), and obesity, the number of affected CKD patients has been increasing rapidly, an estimated almost 850 million worldwide^1^. The amount of creatinine in the blood, calculated by eGFR (estimated glomerular filtration rate), which is considered normal or as healthy kidney if it is within 75-100 ml/min in young adults. However, the range decreases gradually with the age. So, chronic kidney disease can be defined as if eGFR<60 ml/min/1.73m^2^ or any markers of kidney damage (i.e. albuminuria), or both, of at least for three months duration^2^.

CKD stages are determined based on eGFR and the level of proteinuria^3^. End-stage kidney disease (ESKD) or End-stage renal disease (ESRD) or Stage G5 kidney failure is defined when eGFR < 15 ml/min/1.73m^2^ and may be treated with Kidney replacement therapy (KRT) i.e. dialysis, or transplantation, or with supportive care^4^. According to a 2017 study, globally about 3.9 million kidney failure patients were treated with KRT, while approximately equivalent number of patients did not receive any KRT^5^. Dialysis is a process to remove waste products (i.e. nitrogenous substances, urea, creatinine) and excess fluid from the blood when the kidneys are not able to do it. Dialysis can be performed in a hospital, a dialysis center, or at home. Without dialysis, in end-stage kidney disease, the toxic build-up of fluid and waste bi-products in the body can lead to serious health implications, including death. Therefore, dialysis enables improved quality of life as well as give the patients time to prepare for a kidney transplant.

Currently two types of dialysis process are being popular; hemodialysis and peritoneal dialysis. While both the process has their advantages and disadvantages, it helps the patients to lead their regular life. But unfortunately, dialysis is very much costly and impose a heavy burden to the patients and their family. Moreover, the scarcity of dialysis center and dialysate makes it harder for the patients in underdeveloped countries in Asia and Africa.

### Gut Microbiota & Chronic Kidney Disease

An estimated 100 trillion microorganisms live in the lower gastrointestinal (GI) tract of human which is ten times greater than the number of cells in a living organism^6^. Among these gut microbiota, the saccharolytic bacteria plays an essential role in regulating intestinal barrier function^7^, promoting immunity^8^, controlling nutritional uptake and metabolis^9,10^, and preventing proliferation of pathogenic bacteria^11,12^.

Dysbiosis is characterized by a disruption to the microbiome resulting in an imbalance in the microbial species, changes in their functional composition and metabolic activities, or a shift in their local distribution^13^. Studies reported that, as a consequence, the number of pathobionts i.e. Proteobacteria and Fusobacteria were increased in abundance where as beneficial bacteria i.e. Firmicutes was decreased^14^.

Recent study revealed that there is a role of gut microbiota in the development and progression of CKD and is bidirectional. The association between gut microbiota and kidney also known as “gut-kidney axis” (Figure 1)^12^. Dysbiosis of gut microbiota increases uremic toxins, such as p-cresyl (PC), p-cresyl sulfate (PCS), p-cresyl-glucuronide (PCG), indole-3 acetic acid (IAA), indoxyl sulfate (IS), trimethylamine N-oxide (TMAO) and oxalic acid(OA)^15,16,17^. Uremic toxins (UTs) are defined as harmful solutes accumulated in the body when the kidneys are unable to perform their function although they could easily be removed by healthy kidneys^18,19^. Uremic toxins damage the intestinal barrier, resulting increased permeability of this toxins and systemic inflammation^20^. These uremic toxins enter the blood circulation through intestinal wall. In CKD, as the kidney is unable to perform its function of removing these endotoxins, they cause renal endothelial dysfunction, fibrosis, and tubular damage, which subsequently accelerate the progression of CKD^21^. The endotoxin PCS and IS are both associated with higher mortality rate in CKD with Cardiovascular diseases (CVD), and the cardiovascular mortality rate is 10 to 30 times higher in dialysis patients than in the general population^22^. Both the toxins are classified as protein bound and cannot be efficiently removed by hemodialysis^23^. So, therapies that can modulate gut microbiota such as use of probiotics or synbiotic has emerged as an alternative adjuvant therapy in end-stage renal diseae, particularly on dialysis patients to remove uremic toxins, slow down the progression of CKD, and reduce the mortality rate with CVD.

**Figure 1:**
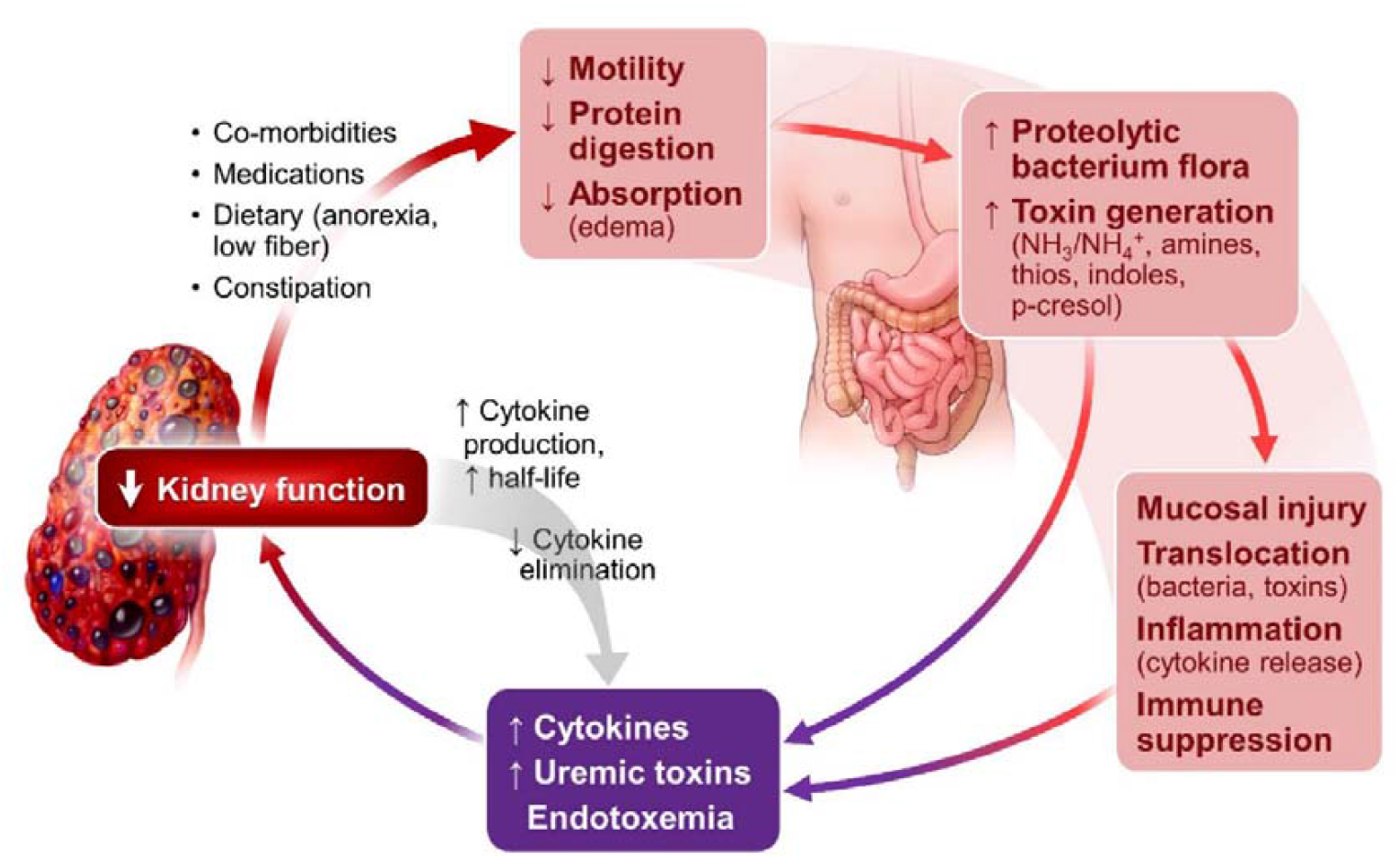
Gut-kidney axis^26^

### Probiotics, Prebiotics, Synbiotic

The term probiotic originated from Greek language which means “for life”. Although the definition of probiotic has changed over time but in 2013, International Scientific Association for probiotics and Prebiotics (ISAPP) defined probiotics as “live microorganisms that, when administered in adequate amounts, confer a health benefit on the host”^24^. Probiotics can be found in our traditional food such as yogurt, kefir, cheese, pickles and in some other foods. Although live microorganisms may be found in many foods and supplements, but only selected species and strains with scientifically proven health benefits should be called probiotics. The most common probiotics that we administer as part of food or a supplement re from *Lactobacillus* and *Bifidobacterium* species. Some of them are *Lactobacillus acidophilus, L. casei, L. paracasei, L. plantarum, L. reuteri, L. rhamnosus, Bifidobacterium longum, B. lactis, B. bifidum, B. infantis* etc. *Streptococcus thermophilus and S. boulardi* are also considered as probiotics^25^.

Along with probiotics, the term prebiotics and synbiotics also come simultaneously. ISAPP defines prebiotics as “a substance that is selectively utilized by host microorganisms conferring a health benefit”^27^. Fructans (fructooligosaccharides (FOS) and galactans (galactooligosaccharides or GOS) are currently dominating the prebiotic category^27^. Prebiotics can be metabolized by bacteria in the gastrointestinal tract to produce beneficial metabolites such as short chain fatty acids (SCFAs)^28^. In 2017, the ISAPP also updated the definition of synbiotic to “a mixture comprising live microorganisms and substrates selectively utilized by most microorganisms that confers a health benefit on the host”^29^. So. synbiotics are a combination of probiotics and prebiotics that are believed to have synergistic effect by inhibiting the growth of pathogenic bacteria and enhancing the growth of beneficial organisms^30^.

The aim of this systematic review is to evaluate the effect of probiotics or synbiotics treatment on dialysis patients in end-stage renal disease.

## METHOD

### Search for Literature and Selection of Studies

This review was carried out according to the Preferred Reporting Items for Systematic Reviews and Meta-Analysis (PRISMA) guidelines (Figure 3). The search for literature was carried out on MEDLINE (accessed via PubMed) database from 2002 to 2023. The searches were carried out using following keyword: (probiotics AND chronic kidney disease; synbiotic AND chronic kidney disease; probiotics AND end-stage renal disease; probiotics AND end-stage kidney disease; gut microbiota AND chronic kidney disease; probiotics AND dialysis; probiotics AND hemodialysis; probiotics AND peritoneal dialysis; Lactobacillus AND chronic kidney disease, symbiotic AND chronic kidney disease).

### Inclusion and Exclusion Criteria

The review included published studies from January, 2002 to March, 2023 in English, designed as randomized clinical trials (RCT), enrolling humans to examine the effect of probiotic/synbiotic on dialysis patients in chronic kidney disease, aged 18 years or older of both sexes. Review papers (systematic or meta-analysis), book chapters, animal trials, and editorials were excluded.

### Data Extraction

Corresponding author searched the database by the keywords described above. Then both authors independently assessed the titles and abstracts found from systematic search according to inclusion and exclusion criteria. Any disagreement between the two reviewers were discussed until consensus was reached.

## RESULT

### Search results

Database search yielded 80 articles, among which 43 were duplicates. After assessing the full-text review according to eligibility criteria and qualitative analysis, fifteen articles were selected for the final review (Figure 2). All the studies in the review were RCTs and performed in different parts of the world.

**Figure 2:**
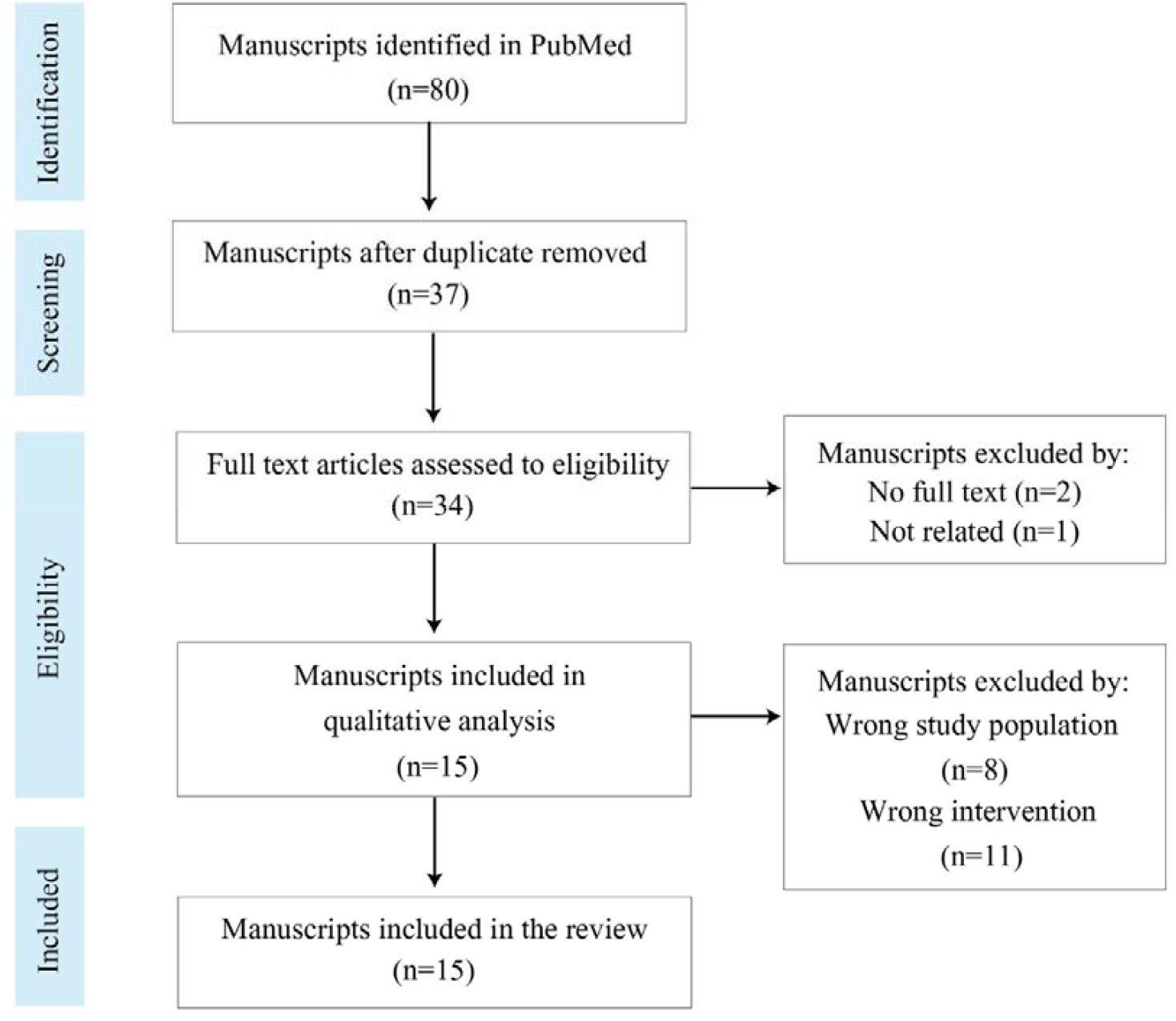
Flow chart showing literature search and screening process using PRISMA guideline

### Characteristics of the articles

Among the fifteen clinical trials, thirteen were performed on hemodialysis patients while two on peritoneal dialysis patients. Sample size was ranged from 9 to 98 patients undergoing dialysis and trial duration varied from four weeks to six months. In most studies, participants were given two or more probiotic combination with prebiotic in either capsule or sachet form. However, few studies used symbiotic gel, also. The quantity of probiotic used and dosage guideline were varied study to study and did not follow any specific standards.

## DISCUSSION

During dialysis, accumulation of uremic toxins, intervention of dialysis catheter and strict dietary restriction could encourage the growth of pathogenic bacteria as well as inhibition of beneficial bacteria, resulting dysbiosis of gut microbiome. Administration of probiotics, single or in combination, could be a viable approach to balance the microbiota balance. Jose Cruz-Mora et al.^31^ found that Bifidobacterium count has increased after the initiation of symbiotic gel to the dialysis patients. The group experimented on eighteen hemodialysis patients of which eight patients received nutritional counselling and a symbiotic gel (containing probiotic combination of *Lactobacillus acidophillus* and *Bifidobacterium bifidum*) along with prebiotic (inulin) and other vitamins while ten patients received nutritional counselling and placebo. Bifidobacterium species produce acetic acid and lactic acid to acidify the gut, resulting prevention of the growth of aerobacteria such as *E. coli* that would produce harmful substances and the growth of pathogenic bacteria^32^. Acetic acid is more effective in reducing the pH in intestine, thus, Bifidobacterium is more effective than Lactobacillus as they cannot produce acetic acid. Another group, Chih-Yu Yang et al.^33^ found that administration of synbiotics (Lactobacillus sp., Bifidobacterium sp., and Streptococcus sp.) could be ameliorated the gut dysbiosis and renal function impairment.

Studies found that, gastrointestinal (GI) symptoms such as anorexia, nausea, vomiting, heartburn, stomachache, bloating, diarrhea and constipation are common in patients with chronic renal disease undergoing dialysis treatment^34,35,36^ although gastro-eosophageal reflux symptoms are more common in peritoneal dialysis (PD) patients compared to hemodialysis (HD) patients or pre-dialytic patients^37^. A possible mechanism behind the increased prevalence of GI symptoms is delayed gastric emptying which is frequent in chronic renal failure^38^ and it further increases during dialysis. Daniela Viramontes-Horner et al.^39^ showed in a double-blind placebo-controlled randomized clinical trial that probiotics significantly reduced the prevalence and monthly episodes of vomit, heartburn, and stomachache as well as a significant decrease in GIS severity in 2 months of treatment. In another study, administration of *Lactobacillus casei* and *Bifidobacterium breve* for 4 weeks in 9 hemodialysis patients resulted in normalization of bowel habits^40^.

**Table 1:** Summary of clinical trials on effect of probiotic in hemodialysis and peritoneal dialysis patients.

| Reference | Study Population | Study Type | Sample Size | Probiotic Type and Intervention | Duration | Conclusion |
| --- | --- | --- | --- | --- | --- | --- |
| Fumio Takayama et al.<sup>32</sup> | Hemodialysis patients | Randomized controlled comparative study | 22 | Bifina (Bifidobacterium longum in gastro-resistant capsule) | 5 weeks | Oral administration of Bifidobacterium to HD patients is effective in reducing the serum levels of indoxyl sulfate by correcting the intestinal microflora. |
| Iwao Nakabayashi et al.<sup>40</sup> | Hemodialysis patients | Interventional study | 9 | Lactobacillus casei and bifidobactrium breve | 4 weeks | Probiotic treatment resulted in normalization of bowel habits and a decreaseof serum p-cresol levels in HD patients |
| Jose Cruz-Mora et al.<sup>31</sup> | Hemodialysis patients | Randomized, double-blind, placebo-controlled clinical trials | 18<br>(Test group, n=8 and Control group, n=10) | Symbiotic gel (Lactobacillus acidophillus and Bifidobacterium bifidum) | 2 months | Short-term symbiotic treatment in patients wirh ESRD can lead to the increase od Bifidobacterium counts, maintaining the intestinal microbial balance. |
| Ranganathan Natarajan et al.<sup>41</sup> | Hemodialysis patients | Randomized, double-blind, place-controlled crossover study | 22 | Renadyl probiotic formulation (Streptococcus thermophilus KB19, Lactobacillus acidophillus KB27, Bifidobacterium longum KB31) | 6 months | Renadyl appeared to be safe to administer to ESRD patients on hemodialysis with encouraging stability in QOL assessment. |
| Daniela Viramontes-Horner et al<sup>39</sup> | Hemodialysis patients | Double-blind, placebo-controlled, randomized clinical trials | 42<br>(Intervention group, n=22 and Control group, n=20) | Symbiotic gel (NutriHealth) (Lactobacillus acidophillus NCFM and Bifidobacterium lactis Bi-07) | 2 months | Administration of asymbiotic gel is a safe and simple way to improve common GIS in dialysis patients. |

**Table 1. Cont.**
| Reference | Study Population | Study Type | Sample Size | Probiotic Type and Intervention | Duration | Conclusion |
| --- | --- | --- | --- | --- | --- | --- |
| Alireza Soleimani et al.<sup>42</sup> | Hemodialysis patients | Parallel randomized double-blind, placebo-controlled clinical trials | 55<br>(Probiotic group, n=28 and Placebo group, n=27) | Lactobacillus acidophilus, Lactobacillus casei, Bifidobacterium bifidum | 12 weeks | Probiotic supplementation among diabetic hemodialysis patients had beneficial effects on parameters of glucose homeostasis, and some biomarkers of inflammation and oxidative stress. |
| Farzad Eidi et al.<sup>43</sup> | Hemodialysis patients | Randomized, place-controlled, double-blind clinical trial | 42<br>Treatment group, n=21 and control group, n=21) | Lactobacillus rhamnosus | 4 weeks | Probiotic could be a promising target in hemodialysis patients with the capability of decreasing serum phenolic uremic toxins. |
| Zahra Shariaty et al.<sup>44</sup> | Hemodialysis patients | Randomized, parallel clinical trials | 36<br>(Intervention group, n=18 and placebo group, n=18) | Lactobacillus acidophilus, Lactobacillus casei, Lactobacillus rhamnosus, Lactobacillus bulgaricus, Bifidobacterium breve, Bifidobacterium longum, Streptococcus thermophilus) | 3 months | Probiotic supplementation decreased Hb fluctuations in hemodialysis patients but did not result in a significant increase in Hb levels. |
| Natalia A. Borges et al.<sup>45</sup> | Hemodialysis patients | Randomized double-blind, placebo-controlled clinical trials | 46<br>(Probiotic group, n=23 and Placebo group, n=23) | Lactobacillus acidophilus, Bifidobacterium longum, Streptococcus thermophilus | 3 months | Probiotic supplementation failed to reduce uremic toxins and inflammatory markers. |
| Maria Teresa Rocchetti et al.<sup>46</sup> | Hemodialysis patients | Randomized placebo-controlled pilot study | 11<br>(Synbiotic group, n=6 and Placebo group, n=5) | NATUREN G (Lactobacillus casei LC4P1, Lactobacillus bulgaricus SP5, Bifidobacterium animalis BLC1 | 8 weeks | Probiotics first line evidence on the synergistic action of gut microbiota modulation aimed at reducing plasma levels of IS and PCS. |

**Table 1. Cont.**
| Reference | Study Population | Study Type | Sample Size | Probiotic Type and Intervention | Duration | Conclusion |
| --- | --- | --- | --- | --- | --- | --- |
| Chih-Yu Yang et al.<sup>33</sup> | Hemodialysis patients | Single center double-blind, placebo-controlled randomized clinical trials | 40 CKD and 22 healthy controls | Synbiotic combination of Lactobacillus sp., Bifidobacterium sp., Streptococcus sp. | 5 weeks | Gut dysbiosis and renal function impairment could be ameliorated by synbiotic treatment. |
| Eunho choi et al.<sup>47</sup> | Hemodialysis patients | Interventional study | 22 | Bifidobacterium longum BORI and Bifidobacterium bifidum BGN4 | 3 months | Probiotic supplementation reduced systemic inflammatory responses in HD patients and this effect was associated with an increase in Tregs and a decrease in proinflammatory monocytes. |
| Aida Lydia et al.<sup>48</sup> | Hemodialysis patients | Double-blind, randomized-controlled clinical trial | 60<br>(Intervention group, n=30 and Control group, n=30) | Synbiotic combination of Lactobacillus acidophilus, Bifidobacterium longum | 2 months | Synbiotic supplement did not lower indoxyl sulfate toxin levels but it had a major effect in improving constipation and quality of life affected by constipation in patients undergoing HD. |
| I.-K. Wang et al.<sup>49</sup> | Peritoneal dialysis patients | Randomized, double-blind placebo-controlled study | 39<br>(Probiotic group, n=21 and Placebo group, n=18) | Bifidobacterium bifidum A218, Bifidobacterium catenulatum A302, Bifidobacterium longum A101, Lactobacillus plantarum A87 | 6 months | Probiotics could significantly reduce the serum levels of endotoxin, TNF-α and IL-6, IL-5, increase the serum levels of IL-10, and preserve residual renal function in PD patients. |
| Yangbin Pan et al.<sup>50</sup> | Peritoneal dialysis patients | Parallel randomized double-blind, clinical trials | 98<br>(Intervention group, n=50 and Control group, n=48) | Bifidobacterium longum, Lactobacillus bulgaricus, Streptococcus thermophilus | 2 months | Probiotics significantly decreases the inflammation marker and health-related quality of life partially improved after probiotic supplementation in patients undergoing PD. |

Many studies have been designed to investigate the effect of probiotics on serum uremic toxins, particularly of Indoxyl Sulfate (IS) and p-cresol sulfate (PCS) as these protein-bound toxins cannot fully excreted by dialysis and are associated with high mortality rate in hemodialysis patients. Fumio Takayama et al.^32^ observed that orally administered Bifidobacterium in a gastro-resistant seamless capsule was effective in reducing the serum level of indoxyl sulfate (IS) in HD patients by correcting the intestinal microflora. In a 2018 study, 21 hemodialysis patients received one capsule containing 1.6*10^7^ CFU of *Lactobacillus rhamnosus* and 21 HD patients received placebo for 4 weeks. Result showed that the mean of serum p-cresol was lower as compared to the baseline (2.68 mg/dL vs 1.23 mg/dL; p=0.034) and a same trend was observed for the serum p-cresol levels phenol (2.10 mg/dL vs 1.01 mg/dL; p = 0.009) but in the placebo group, the serum concentrations of the toxins did not decrease statistically^43^. Eunho choi et al.^47^ found that probiotics supplementation were able to decrease proinflammatory monocytes which reduced the systemic inflammatory response in HD patients. In another study with peritoneal dialysis patients, I.-K. Wang et al.^49^ observed that levels of serum TNF-α , IL-5, IL-6 and endotoxin significantly decreased after six months of probiotic treatment, while levels of serum IL-10 significantly increased. Yangbin Pan et al.^50^ also found that administration of probiotics significantly decreased serum c-reactive protein and IL-6 and increased the serum albumin levels in PD patients. On the other hand, Natalia A. Borges et al.^45^ showed that probiotic supplementation (Lactobacillus acidophillus, Bifidobacterium longum, Streptococcus thermophilus) failed to reduce uremic toxins and inflammatory markers HD patients.

Another group, Alireza Soleimani et al.^42^ observed the effect of probiotics on diabetic hemodialysis patients. Probiotics supplements of Lactobacillus acidophillus, Lactobacillus casei and Bifidobacterium bifidum compared with placebo showed significantly decreased fasting plasma glucose (-22.0 vs +6.6 mg/dL), serum insulin (-6.4 vs +2.3 µIU/ml), and HbA1c (-0.4 vs -0.1%), and improved quantitative insulin sensitivity check index (+0.03 vs -0.02) in 12 weeks treatment. Moreover, they found that probiotic supplementation significantly reduced serum c-reactive protein (-1933 vs. +252 ng/mL), plasma malondialdehyde (-0.3 vs. +1.0 µmol/l), subjective global assessment score s (-0.7 vs. +0.7) and significantly increased plasma total antioxidant capacity (+15 vs. -88 mmol/l). In another study, the researchers found that probiotics decreased Hb fluctuations in HD patients but could not able to increase the Hb level significantly.

### Limitations of the Reviewed Studies

There are several noteworthy limitations in our reviewed studies. Firstly, the sample sizes of the studies were small and study durations were not long enough to detect a significant change. Secondly, the diversity of probiotics and combination criteria, dose and intervention period made it harder to compare among them. Thirdly, scarcity of clinical data on the use of probiotics in PD patients. Additionally, the formulation of probiotics may also play an important role on their efficacy as they need to pass the harsh acidic condition before reaching intestine.

## CONCLUSION

In conclusion, our review demonstrates administration of probiotics, as single or in combination have potential beneficial effects on dialysis patients in end-stage kidney disease. Probiotics may exert positive effect against gut dysbiosis, GI symptoms, uremic toxins, inflammation and overall quality of life of dialysis patients. Future large-scale clinical studies using a larger sample size with longer study duration are required to elucidate the benefits of probiotics on dialysis patients, particularly in PD patients.

## Data Availability

All data produced in the present study are available upon reasonable request to the authors.

## Author’s contribution

All authors had access to the data and a role in writing the manuscript.

